# Detection of parasite-derived transrenal DNA for the diagnosis of chronic *Trypanosoma cruzi* infection

**DOI:** 10.1101/19010934

**Authors:** Ana G. Madrigal, Rachel Marcus, Robert Gilman, Alan L. Scott, Clive Shiff

## Abstract

Chagas disease, caused by the protozoan parasite *Trypanosoma cruzi*, is a potentially life-threatening infection endemic to Latin America that has emerged as a global public health issue due to globalization and emigration patterns. Diagnosis of *T. cruzi* infection is complex, especially for the chronic phase of the disease that is characterized by a low to moderate burden of the difficult to detect, tissue-dwelling, intracellular form of the parasite. Diagnosis relies on a multistep indirect serological detection approach that requires positive results in at least two independent anti-*T. cruzi* antibody tests. With no gold standard diagnostic method for chronic *T. cruzi*, new approaches are needed that can more directly test for the presence of the parasite. Here, we report on of the potential utility of a noninvasive diagnostic approach that specifically detects *T. cruzi*-derived cell-free repeat DNA in the urine of patients who are both serologically positive and negative.

## Introduction

Chagas disease is a vector borne, neglected tropical infection endemic to Latin America. The etiological agent of Chagas disease, the hemoflagellate *Trypanosome cruzi*, is maintained in the environment by a zoonotic cycle that includes the triatomid vector and a variety of mammalian species. Recently, Chagas disease has emerged as a global public health issue due to globalization and the changing patterns of migration (1, 2); with approximately 8 million people infected worldwide and at least 100 million at risk of infection (3). This complex disease involves an acute and chronic phase that can cause heart and gastrointestinal issues depending on disease progression. While Chagas disease was formally described by Carlos Chagas 110 years ago, there is still no single test for definitive diagnosis of the chronic form of *T. cruzi* infection and the estimated index of under diagnosis remains ∼95% (3).

In the acute phase (6-8 weeks post-infection), extracellular trypomastigotes are found in the blood and, depending on the intensity of infection, can be readily detected through direct microscopic visualization (3). Following the acute phase, around 70% of patients clear the infection. The remaining patients transition into the chronic manifestation of the infection, which can last 10-30 years (3) and where the non-circulating, difficult to detect intracellular amastigote is the dominate form of the parasite. Diagnostics for chronic infection is based on a patchwork of patient’s history, clinical findings and the results of multiple serological tests designed to detect parasite antigen-specific IgG antibodies (4). Confirmatory diagnosis requires two different serological tests, with a third test for confirmation when results are incongruous. When infection is suspected, ECG’s, X-rays, endoscopy, and other tests are run to assess the presence of Chagas disease-associated cardiomyopathy, megacolon, and megaesophagus (5). Importantly, serological tests vary in sensitivity by geographical regions (6), further complicating diagnosis.

These factors result in a serious under-diagnosis of *T. cruzi* infections in both endemic and non-endemic areas.

Trypanomastid genomic DNA is complex with ∼50% of the genome consisting of repeated sequences (7). In addition to the nuclear genome, the kinetoplast genome, which represents ∼30% of the total DNA of the cell, is also highly repetitive with thousands of 1.4 kb minicircles and dozens of ∼25 Mb maxicircles (8). The repetitive nature of the genome provides multiple targets for PCR-based-detection approaches. Two of these targets have been termed satellite DNA (or Sat-DNA) and kinetoplast DNA (or K-DNA). The Sat-DNA designated TCZ is composed of ∼120,000 copies of a tandemly repeated 195 bp sequence of non-coding DNA (23). TCZ represents ∼10% of the parasites’ total DNA (9). The K-DNA minicircles each of contain four regions of highly conserved sequence. The abundance of these repeats has made them the focus of PCR-based approaches to detect the parasite DNA in blood and tissues (10).

The use of biomarkers in blood and other bodily fluids has gained wide use in the clinic. Of note, the use of cell-free DNA is being applied as a biomarker for cancer, prenatal diagnosis and in infectious diseases (11-13). While most methods use blood, cell-free DNA is also readily detected in urine (13-16) and other biological fluids (17, 18). The detection of cell-free DNA has been demonstrated to be of utility for the diagnosis of malaria, trypanosomiasis, leishmaniasis, schistosomiasis, strongyloidiasis, and filariasis (reviewed in (12)). In the blood, cell-free DNA is degraded by endogenous nucleases (19) with a majority of the degradation products being cleared by the liver and kidney (20). Further, it has been demonstrated that a proportion of the cell-free DNA fragments identified in the circulation can pass through the glomerular barrier and appear as 100-300 pb transrenal DNAs in the urine (14).

In the work presented here, we focused on determining the potential utility of detecting parasite-derived transrenal DNAs by PCR to aid in the diagnosis of patients with chronic *T. cruzi* infection. The results demonstrated that both *T. cruzi* Sat-DNA repeat and minicircle K-DNA can be detected in the urine of both serology positive and serology negative patients and that urine-based diagnosis has the potential to be a useful alternative approach to supplement current diagnostic methods.

## Materials and Methods

### Sample collection

A total of 36 urine and 34 blood samples were collected following informed consent from patients participating in a study looking at the cardiac complications associated with chronic Chagas disease directed by Dr. Robert Gilman (Department of International Health, BSPH, JHU) and Dr. Rachel Marcus (MedStar Health & Vascular Institute, Washington, DC).

The study was part of a program to study the seroprevalence and characterization of Chagas Disease cardiomyopathology among Latin Americans living in Fairfax County, VA. This was approved by Johns Hopkins Bloomberg School of Public Health IRB No: 00006713 amended on 5.March 2018 to include this work. All participants fully understood the objective and participated willingly. Participants provided ∼45 mL of urine to which EDTA was added to 40 mM (pH 8). The fresh urine was processed within 7 hours of collection by filtering through a filter paper cone (12.5 cm Whatman No. 3), dried overnight, and stored individually with a desiccant in plastic sleeves at 4°C. Urine samples obtained from ten individuals with no clinical history and who were serologically negative were used as negative controls.

### Serology

Serum samples were collected to assess the presence of anti-*T. cruzi* antibodies using the following serological tests: Chagas Detect Plus (InBios, Seattle, WA), Hemagen Chagas Kit (Hemagen Diagnostics, Inc. Columbia, MD), Chagatest EIA Recombinant V3.0 (Wiener Laboratorios S.A.I.C., Rosario, Argentina), BIOZIMA Chagas Kit (Polychaco, S.A.I.C., Buenos Aries, Argentina), and immunofluorescent detection for the presence of anti-epimastigote antigens.

### Isolation of transrenal DNAs

Ten 6.35 mm discs were punched from the inner quadrant of the dried filter and place in a 1.5 ml tube. The discs were covered with 400 µL of nuclease-free water, incubated at 95°C for 10 minutes, and then gently agitated overnight in a rotating water bath at room temperature (∼18 hours). After centrifuging at 4,000 rpm for 5 minutes, the transrenal DNA was isolated using QIAmp DNA Blood Mini Kit (Qiagen, MD) following the manufacturer’s protocol. DNA concentrations were measured using the Nano Drop ND-1000 spectrophotometer (NanoDrop Technologies, Wilmington, DE) and samples were stored at - 20°C.

### PCR to detect *T. cruzi* transrenal DNAs

Three primer sets were employed to detect two distinct *T. cruzi* repeat families. Two primer sets were used to detect K-DNA minicircle repeats. Primers designated minicon-1 (M1) and minicon-2 (M2) (**Figure 1**) amplified a 113 bp product from the *T. cruzi* K-DNA. The PCR reaction was prepared to a final volume of 15 µL containing 7.5 µL of PCR Master Mix (New England Biolabs, Ipswich, MA), 0.75 µL of each primer (10 µM), 2 µL of MgCl_2_ (25 mM), 2 µL of DNAs-free water and 2 μL of transrenal DNA template (1-3 ng/μL).

**Figure 1.**
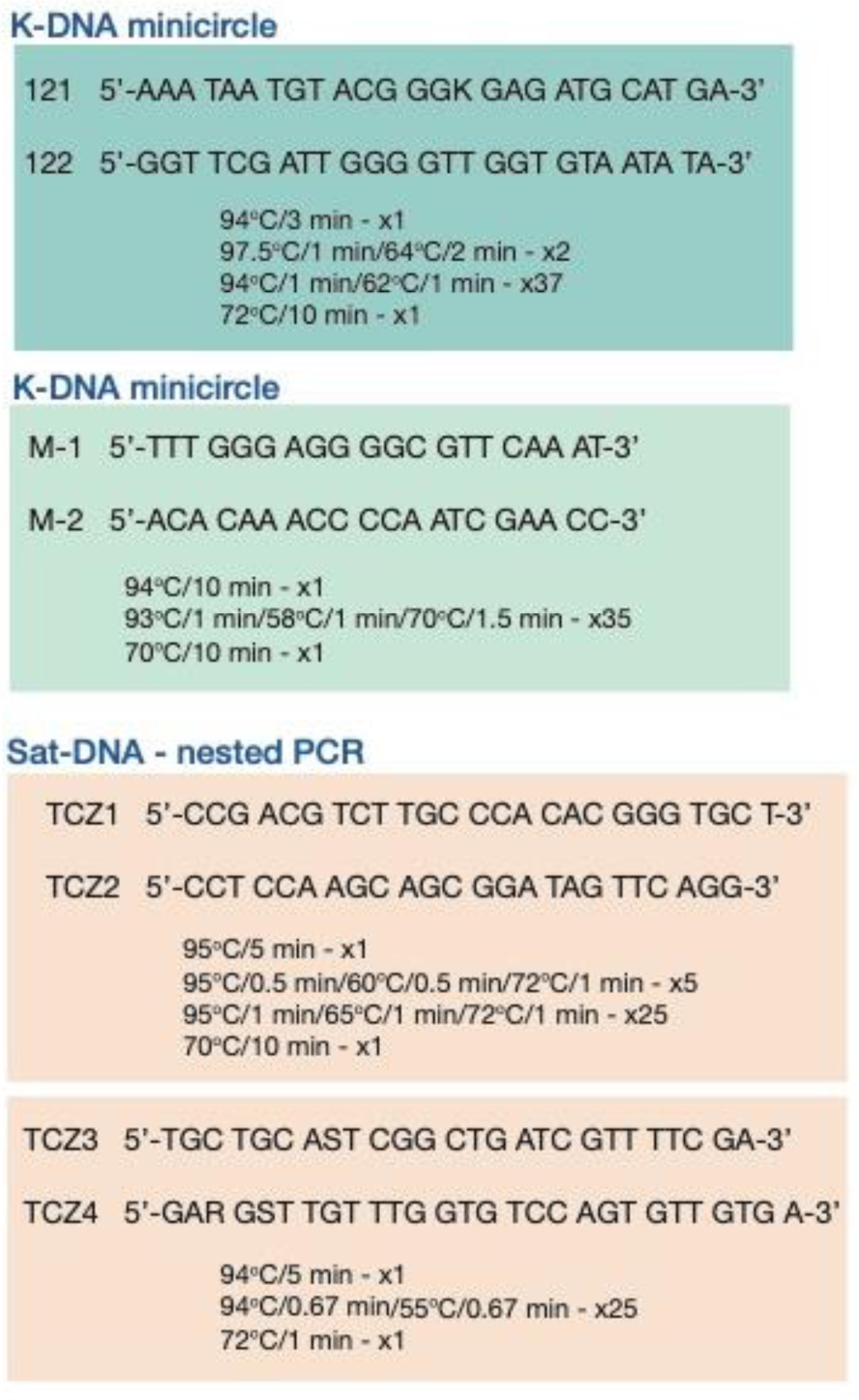
Summary of primers targeting *T. cruzi* K-DNA minicircle and SAT-DNA repeats and the amplification conditions used for each of the primer sets. Primer set 121/122 is after Shijman et al. (12). The TCZ primer sets nPCR are after are after Martins et al. (21). For TCZ3 and TCZ4 primer sequences, a S indicates a G or C and a R indicates an A or G at these positions.

Primers 121-122 (**Figure 1**), targeting the K-DNA minicircle variable domain amplified a 330 bp product Shijman et al. (12). Amplification was performed at 15 µL final volume, containing 7.5 µL of PCR Master Mix (New England Biolabs, Ipswich, MA), 0.75 µL of each primer (10 µM), 0.75 µL of MgCl_2_ (25 mM), 3.25 µL of DNAs-free water, and 2 μL of transrenal DNA template (1-3 ng/μL).

The third set of primers used a nested PCR (nPCR) approach based on Martins et al. (21) employing primer sets TCZ1/TCZ2 and TCZ3/TCZ4 (**Figure 1**) in a two-step PCR that produced a 149 bp product from the *T. cruzi* Sat-DNA repeats. For the first step amplification was prepped for a total reaction volume of 15 µL containing 7.5 µL of PCR Master Mix (New England Biolabs, Ipswich, MA), 0.75 µL of each primer TCZ1/TCZ2 (10 µM), 1.5 µL of MgCl_2_ (25 mM), 2.5 µL of DNAs-free water, and 2 μL of transrenal DNA template (1-3 ng/μL). For the second step, 7 µL of previous PCR product was used in a final volume of 20 µL, containing 7.5 µL of PCR Master Mix (New England Biolabs, Ipswich, MA), 0.75 µL of each primer TCZ3/TCZ4 (10 µM), 1.5 µL of MgCl_2_ (25 mM), and 2.5 µL of DNAs-free water. A summary of the primer sequences and amplification conditions are outlined in **Figure 1**.

Each PCR run included a positive control composed of *T. cruzi* genomic DNA, five negative urine controls and a water-only control. Each sample was assayed in duplicate and amplifications were conducted in a SimpliAmp™ Thermal Cycler (Life Technologies™). The amplicons were initially assessed by size on a 2% agarose gel. All bands were extracted and confirmed by sequencing as *T. cruzi* repeat DNA.

## Results

Urine samples were obtained from volunteers who originated in areas endemic for *T. cruzi* but now reside in the mid-Atlantic region of the U.S. These volunteers visited the clinic for the diagnosis and treatment of cardiovascular conditions, some of which are associated with chronic Chagas disease. Of the 36 volunteers who provided urine samples, 9 (25%) were male, 25 (69.4%) were female, and 2 (5.5%) did not respond. The overall mean age of the volunteers was 42.1 (± SD 11.8). More than half of the participants (19; 52.8%) were between 40-59 years old and 30.6% were between 18-39 years old. A majority of the participants originated from either El Salvador (18, 50%) or Bolivia (11, 30.6%), with two from Argentina, two from Guatemala, one from Costa Rica, and one from Peru (**Table 1**). One individual did not provide their country of origin (NP, not provided). None of the volunteers who originated from *T. cruzi*-endemic regions showed overt signs of disease.

**Table 1.** Results from serological-based detection of circulating T. cruzi antigen; and PCR-based detection of T. cruzi transrenal cfDNA from urine samples obtained from patients.

| Sample | Origin | Serology | nPCR | Minicon | 121-122 |
| --- | --- | --- | --- | --- | --- |
| 1 | ES | + | + | + | + |
| 2 | A | - | + | + | - |
| 3 | B | - | + | + | - |
| 4 | B | + | + | - | - |
| 5 | ES | - | + | - | - |
| 6 | A | - | + | - | - |
| 7 | B | + | + | - | - |
| 8 | B | + | - | + | - |
| 9 | B | - | - | + | - |
| 10 | G | - | - | + | - |
| 11 | B | - | - | + | - |
| 12 | ES | - | - | + | - |
| 13 | NP | NA | - | + | - |
| 14 | B | + | - | - | - |
| 15 | ES | + | - | - | - |
| 16 | ES | + | - | - | - |
| 17 | ES | + | - | - | - |
| 18 | ES | + | - | - | - |
| 19 | B | + | - | - | - |
| 20 | B | + | - | - | - |
| 21 | B | + | - | - | - |
| 22 | ES | + | - | - | - |
| 23 | ES | + | - | - | - |
| 24 | ES | + | - | - | - |
| 25 | ES | + | - | - | - |
| 26 | ES | - | - | - | - |
| 27 | ES | - | - | - | - |
| 28 | G | - | - | - | - |
| 29 | CR | - | - | - | - |
| 30 | ES | - | - | - | - |
| 31 | P | - | - | - | - |
| 32 | ES | - | - | - | - |
| 33 | ES | - | - | - | - |
| 34 | B | NA | - | - | - |
| 35 | ES | - | - | - | - |
| 36 | ES | - | - | - | - |

A = Argentina; B = Bolivia; CR = Costa Rica; ES = El Salvador;
G = Guatemala; P = Peru; NP = Not Provided.
+ and - indicate that the outcome of the assay was positive or negative, respectively; NA = Not Applicable; cf-DNA = cell-free DNA.

Approximately half of the volunteers (16/36) were considered positive for exposure to *T. cruzi* by serology (**Table 1**). In order to be designated positive, a serum sample had to give a positive result in at least two of the five antibody-based diagnostic assays (see Methods section).

*T. cruzi* transrenal DNA was confirmed in 36% (13/36) of the urine samples. Of the 13 urine samples that were positive for *T. cruzi* transrenal DNA, four were obtained from individuals who also tested positive for antibodies. There was no apparent pattern for detection of transrenal DNA based on the country of origin or the sex of the volunteer.

The Minicon primers that targeted the K-DNA conserved domain amplified a 113 bp fragment from nine samples. The nPCR approach targeting Sat-DNA amplified a 149 bp fragment from seven samples. Primers 121-122 that targeted the K-DNA variable domain amplified a 330 bp product from only a single sample. Of the 13 samples that tested positive for *T. cruzi* transrenal DNA, three were concordantly positive between the nPCR and Minicon approaches. Only one sample was positive for all three *T. cruzi* transrenal DNA amplicons.

## Discussion

Recently, interest has increased in using urine as a ‘liquid biopsy’ to diagnose and monitor the efficacy of intervention strategies for viral, bacterial and parasitic pathogens (22-24). While some of these tests are designed to detect pathogen-derived proteins and glycoproteins, many focus on the detection of pathogen-derived transrenal DNA. The advantages of transrenal DNA-based diagnosis of infectious diseases include (a) urine collection is non-invasive and abundant (especially important for problematic patients such as neonates), (b) urine is easy and cheap to collect and process (important for field work), (c) diagnosis does not depend on the stage of the parasite or the site of infection, (d) the use of urine circumvents the multiple PCR inhibitors that are present in the serum (25).

The recent introduction of PCR to detect *T. cruzi* cell-free DNA in blood specimens holds promise as a sensitive and specific alternative to serology (26-28). While PCR assays from blood are sensitive for diagnosis of acute infections, the blood-based assays are much less effective in the context of chronic infections (10). A number of issues, including low levels and intermittent release of *T. cruzi* cell-free DNA, could limit the utility of the blood-based PCR assays. The feasibility to using parasite transrenal DNA in urine to detect *T. cruzi* infection was demonstrated by Castro-Sequen and colleagues (29) who, in a guinea pig model, were able to PCR amplify parasite kinetoplast and nuclear transrenal DNA. We have recently extended this to human urine samples. To our knowledge, our results represent the first demonstration of parasite transrenal DNA from *T. cruzi*-infected humans.

In the blood, cell-free DNA is degraded by endogenous nucleases (19) with a majority of the degradation products being cleared by the liver and kidney (20). Further, it has been demonstrated that cell-free DNA fragments identified in the circulation can pass through the glomerular barrier and appear as transrenal DNA in the urine (14). Reports have shown that transrenal DNAs represent fragments that range between 150–300 bp (30). This small size range needs to be taken into consideration in the design of detection assays. Indeed, in the work presented here, only the Minicon primers that amplified a 113 bp product from the minicircle K-DNA was sensitive in our assay. In contrast, the 121-122 primers, that amplifies a 330 bp amplicon, was significantly less sensitive for the detection of transrenal fragments of minicircle K-DNA with only one sample giving a positive result.

To address the fact that *T. cruz*i persists at low-levels in the tissues as amastigotes and only intermittently in the blood during chronic infection, a number of indirect serological methods have been developed to identify infected individuals. The current WHO-recommended standard for serological detection of *T. cruzi* infection is the use of “two conventional tests based on different principles and detecting different antigens” (31). These most widely used versions of these tests, a number of which were used in this study, measure anti-*T. cruzi* IgG by ELISA, IFA, RIA, Western Blots, and other methods (3, 32). While these tests are reported to have sensitivity and specificity levels between 90-98%, a recent analysis concluded that the serological tests “appear less accurate than previously thought” and that caution should be exercised when using these tests for the identification of individuals with chronic infection (32, 33). Although multiple issues contribute to the uncertainty in the interpretation of anti-*T. cruzi* serology results, the lack of a ‘gold standard’ method or set of reference reagents that can be used to calibrate the various assays and the inability to determine if the antibodies represent on-going or past exposure to the parasite limit the diagnostic utility of these tests.

In the results presented here only 30% of the samples that tested positive for one or more of the *T. cruzi* transrenal repeat sequences were derived from individuals who also meet the criteria to be designated positive by serology. Since the detection of parasite-derived DNA is a direct measure of the ongoing infection, this indicates that, in the context of chronic infection, serology might be seriously underestimating the levels of infection.

Conversely, 12/16 volunteers who tested positive by serology provided urine samples that were negative for transrenal DNAs from both of the parasite repeats. The presence of anti-*T. cruzi* antibodies in the serum of these individuals could reflect previous exposure(s) to the parasite that were subsequently cleared thus accounting for the lack of parasite-derived transrenal DNAs. It is also possible that at least some of these seropositive volunteers are chronically infected and have

*T. cruzi* transrenal DNA in their urine but the parasite-derived repeat DNA that is too low to detect under the conditions employed here. Alternatively, the parasite-derived transrenal DNA might originate from other areas of the parasite’s genome so that the primers used here would miss these patients. Studies designed to identify additional common parasite*-*derived transrenal DNAs would be important for the development of multiplex approach that could enhance the utility of using urine-based analysis as a main-line tool for the clinical and epidemiological monitoring of chronic *T. cruzi* infections.

While further studies with a larger population are needed for improved primer development, the detection of *T. cruzi* DNA in urine has the potential to provide a novel non-invasive and direct diagnostic method for Chagas disease that can be employed to improve chronic phase detection, treatment-monitoring, early detection in congenital disease cases and the management of immunosuppressed patient.

## Data Availability

all data are available in the form a thesis written by the first author

## Acknowledgements

The present study was supported by the Department of Molecular Microbiology and Immunology [Bloomberg School of Public Health, Johns Hopkins University, Baltimore, Maryland]. We thank Dr. Monica Mugnier for her insights and guidance in understanding sequencing results and parameters. We also thank the Zavala lab for allowing the use of their equipment. Finally, we thank Yagahira Castro for her aid in sample collection.

## References

1. Lee, B. Y., K. M. Bacon, M. E. Bottazzi, and P. J. Hotez. 2013. Global economic burden of Chagas disease: a computational simulation model. The Lancet infectious diseases 13: 342–348.

2. Perez, C. J., A. J. Lymbery, and R. C. A. Thompson. 2015. Reactivation of Chagas Disease: Implications for Global Health. Trends Parasitol 31: 595–603.

3. Perez-Molina, J. A., and I. Molina. 2018. Chagas disease. Lancet.

4. Basile, L., J. M. Jansa, Y. Carlier, D. D. Salamanca, A. Angheben, A. Bartoloni, J. Seixas, T. Van Gool, C. Canavate, M. Flores-Chavez, Y. Jackson, P. L. Chiodini, P. Albajar-Vinas, and D. Working Group on Chagas. 2011. Chagas disease in European countries: the challenge of a surveillance system. Euro surveillance : bulletin Europeen sur les maladies transmissibles = European communicable disease bulletin 16.

5. Rassi, A., Jr., A. Rassi, and J. A. Marin-Neto. 2010. Chagas disease. Lancet 375: 1388–1402.

6. Martin, D. L., M. Marks, G. Galdos-Cardenas, R. H. Gilman, B. Goodhew, L. Ferrufino, A. Halperin, G. Sanchez, M. Verastegui, P. Escalante, C. Naquira, M. Z. Levy, and C. Bern. 2014. Regional variation in the correlation of antibody and T-cell responses to Trypanosoma cruzi. Am J Trop Med Hyg 90: 1074–1081.

7. El-Sayed, N. M., P. J. Myler, D. C. Bartholomeu, D. Nilsson, G. Aggarwal, A. N. Tran, E. Ghedin, E. Worthey, A. L. Delcher, G. Blandin, S. J. Westenberger, E. Caler, G. C. Cerqueira, C. Branche, B. Haas, A. Anupama, E. Arner, L. Aslund, P. Attipoe, E. Bontempi, F. Bringaud, P. Burton, E. Cadag, D. A. Campbell, M. Carrington, J. Crabtree, H. Darban, J. F. da Silveira, P. de Jong, K. Edwards, P. T. Englund, G. Fazelina, T. Feldblyum, M. Ferella, A. C. Frasch, K. Gull, D. Horn, L. Hou, Y. Huang, E. Kindlund, M. Klingbeil, S. Kluge, H. Koo, D. Lacerda, M. J. Levin, H. Lorenzi, T. Louie, C. R. Machado, R. McCulloch, A. McKenna, Y. Mizuno, J. C. Mottram, S. Nelson, S. Ochaya, K. Osoegawa, G. Pai, M. Parsons, M. Pentony, U. Pettersson, M. Pop, J. L. Ramirez, J. Rinta, L. Robertson, S. L. Salzberg, D. O. Sanchez, A. Seyler, R. Sharma, J. Shetty, A. J. Simpson, E. Sisk, M. T. Tammi, R. Tarleton, S. Teixeira, S. Van Aken, C. Vogt, P. N. Ward, B. Wickstead, J. Wortman, O. White, C. M. Fraser, K. D. Stuart, and B. Andersson. 2005. The genome sequence of Trypanosoma cruzi, etiologic agent of Chagas disease. Science 309: 409–415.

8. Minning, T. A., D. B. Weatherly, S. Flibotte, and R. L. Tarleton. 2011. Widespread, focal copy number variations (CNV) and whole chromosome aneuploidies in Trypanosoma cruzi strains revealed by array comparative genomic hybridization. BMC Genomics 12: 139.

9. Vargas, N., A. Pedroso, and B. Zingales. 2004. Chromosomal polymorphism, gene synteny and genome size in T. cruzi I and T. cruzi II groups. Mol Biochem Parasitol 138: 131–141.

10. Schijman, A. G., M. Bisio, L. Orellana, M. Sued, T. Duffy, A. M. M. Jaramillo, C. Cura, F. Auter, V. Veron, Y. Qvarnstrom, S. Deborggraeve, G. Hijar, I. Zulantay, R. H. Lucero, E. Velazquez, T. Tellez, Z. S. Leon, L. Galvao, D. Nolder, M. M. Rumi, J. E. Levi, J. D. Ramirez, P. Zorrilla, M. Flores, M. I. Jercic, G. Crisante, N. Anez, A. M. Castro, C. I. Gonzalez, K. A. Viana, P. Yachelini, F. Torrico, C. Robello, P. Diosque, O. T. Chavez, C. Aznar, G. Russomando, P. Buscher, A. Assal, F. Guhl, S. S. Estani, A. DaSilva, C. Britto, A. Luquetti, and J. Ladzins. 2011. International Study to Evaluate PCR Methods for Detection of Trypanosoma cruzi DNA in Blood Samples from Chagas Disease Patients. Plos Neglect Trop D 5.

11. Wagner, J. 2012. Free DNA--new potential analyte in clinical laboratory diagnostics? Biochemia medica 22: 24–38.

12. Weerakoon, K. G., and D. P. McManus. 2016. Cell-Free DNA as a Diagnostic Tool for Human Parasitic Infections. Trends Parasitol 32: 378–391.

13. Yu, J., G. Gu, and S. Ju. 2014. Recent advances in clinical applications of circulating cell-free DNA integrity. Laboratory medicine 45: 6–11.

14. Botezatu, I., O. Serdyuk, G. Potapova, V. Shelepov, R. Alechina, Y. Molyaka, V. Ananev, I. Bazin, Garin, M. Narimanov, V. Knysh, H. Melkonyan, S. Umansky, and A. Lichtenstein. 2000. Genetic analysis of DNA excreted in urine: a new approach for detecting specific genomic DNA sequences from cells dying in an organism. Clinical chemistry 46: 1078–1084.

15. Ibironke, O. A., A. E. Phillips, A. Garba, S. M. Lamine, and C. Shiff. 2011. Diagnosis of Schistosoma haematobium by detection of specific DNA fragments from filtered urine samples. Am J Trop Med Hyg 84: 998–1001.

16. Lodh, N., R. Caro, S. Sofer, A. Scott, A. Krolewiecki, and C. Shiff. 2016. Diagnosis of Strongyloides stercoralis: Detection of parasite-derived DNA in urine. Acta tropica 163: 9–13.

17. Jiang, W. W., B. Masayesva, M. Zahurak, A. L. Carvalho, E. Rosenbaum, E. Mambo, S. Zhou, K. Minhas, N. Benoit, W. H. Westra, A. Alberg, D. Sidransky, W. Koch, and J. Califano. 2005. Increased mitochondrial DNA content in saliva associated with head and neck cancer. Clin Cancer Res 11: 2486–2491.

18. van der Drift, M. A., C. F. Prinsen, B. E. Hol, A. S. Bolijn, M. A. Jeunink, P. N. Dekhuijzen, and F. B. Thunnissen. 2008. Can free DNA be detected in sputum of lung cancer patients? Lung cancer 61: 385–390.

19. Suzuki, N., A. Kamataki, J. Yamaki, and Y. Homma. 2008. Characterization of circulating DNA in healthy human plasma. Clinica chimica acta; international journal of clinical chemistry 387: 55–58.

20. Tsumita, T., and M. Iwanaga. 1963. Fate of injected deoxyribonucleic acid in mice. Nature 198: 1088–1089.

21. Martins, M., M. B. Pereira, J. J. G. Ferreira, A. O. Franca, M. C. Cominetti, E. C. Ferreira, M. Dorval, C. L. Rossi, S. B. Mazon, E. A. de Almeida, S. C. B. Costa, and G. E. B. Marcon. 2018. Serological and molecular inquiry of Chagas disease in an Afro-descendant settlement in Mato Grosso do Sul State, Brazil. PLoS One 13: e0189448.

22. Lu, T., and J. Li. 2017. Clinical applications of urinary cell-free DNA in cancer: current insights and promising future. American journal of cancer research 7: 2318–2332.

23. Wang, J., S. Chang, G. Li, and Y. Sun. 2017. Application of liquid biopsy in precision medicine: opportunities and challenges. Frontiers of medicine 11: 522–527.

24. Zhang, Y. C., Q. Zhou, and Y. L. Wu. 2017. The emerging roles of NGS-based liquid biopsy in non-small cell lung cancer. Journal of hematology & oncology 10: 167.

25. Su, Y. H., M. Wang, D. E. Brenner, P. A. Norton, and T. M. Block. 2008. Detection of mutated Kras DNA in urine, plasma, and serum of patients with colorectal carcinoma or adenomatous polyps. Ann N Y Acad Sci 1137: 197–206.

26. Duffy, T., M. Bisio, J. Altcheh, J. M. Burgos, M. Diez, M. J. Levin, R. R. Favaloro, H. Freilij, and A. G. Schijman. 2009. Accurate real-time PCR strategy for monitoring bloodstream parasitic loads in chagas disease patients. PLoS Negl Trop Dis 3: e419.

27. Ramirez, J. C., C. I. Cura, O. da Cruz Moreira, E. Lages-Silva, N. Juiz, E. Velazquez, J. D. Ramirez, A. Alberti, P. Pavia, M. D. Flores-Chavez, A. Munoz-Calderon, D. Perez-Morales, J. Santalla, P. Marcos da Matta Guedes, J. Peneau, P. Marcet, C. Padilla, D. Cruz-Robles, E. Valencia, G. E. Crisante, G. Greif, I. Zulantay, J. A. Costales, M. Alvarez-Martinez, N. E. Martinez, R. Villarroel, S. Villarroel, Z. Sanchez, M. Bisio, R. Parrado, L. Maria da Cunha Galvao, A. C. Jacome da Camara, B. Espinoza, B. Alarcon de Noya, C. Puerta, A. Riarte, P. Diosque, S. Sosa-Estani, F. Guhl, I. Ribeiro, C. Aznar, C. Britto, Z. E. Yadon, and A. G. Schijman. 2015. Analytical Validation of Quantitative Real-Time PCR Methods for Quantification of Trypanosoma cruzi DNA in Blood Samples from Chagas Disease Patients. The Journal of molecular diagnostics : JMD 17: 605–615.

28. Schijman, A. G., J. Altcheh, J. M. Burgos, M. Biancardi, M. Bisio, M. J. Levin, and H. Freilij. 2003. Aetiological treatment of congenital Chagas’ disease diagnosed and monitored by the polymerase chain reaction. The Journal of antimicrobial chemotherapy 52: 441–449.

29. Castro-Sesquen, Y. E., R. H. Gilman, V. Yauri, J. Cok, N. Angulo, H. Escalante, and C. Bern. 2013. Detection of soluble antigen and DNA of Trypanosoma cruzi in urine is independent of renal injury in the guinea pig model. PLoS One 8: e58480.

30. Su, Y. H., M. Wang, D. E. Brenner, A. Ng, H. Melkonyan, S. Umansky, S. Syngal, and T. M. Block. 2004. Human urine contains small, 150 to 250 nucleotide-sized, soluble DNA derived from the circulation and may be useful in the detection of colorectal cancer. The Journal of molecular diagnostics : JMD 6: 101–107.

31. WHO. 2010. Chagas disease (American trypanosomiasis): Fact Sheet 340.

32. Afonso, A. M., M. H. Ebell, and R. L. Tarleton. 2012. A systematic review of high quality diagnostic tests for Chagas disease. PLoS Negl Trop Dis 6: e1881.

33. Santos, F. L., W. V. de Souza, S. Barros Mda, M. Nakazawa, M. A. Krieger, and M. Gomes Yde. 2016. Chronic Chagas Disease Diagnosis: A Comparative Performance of Commercial Enzyme Immunoassay Tests. Am J Trop Med Hyg 94: 1034–1039.

